# When AI Meets the FDA: An Evaluation of Large Language Models Performance in Regulatory and Clinical Trial Data Extraction, Synthesis, and Analysis

**DOI:** 10.64898/2025.12.22.25342875

**Authors:** Khulud Bukhari, Rosa Rodriguez-Monguio, Beatriz Lopez-Bermudez, Jason Yamaki, Lawrence M. Brown, Richard Beuttler, Jasmine Chiat Ling Ong, Enrique Seoane-Vazquez

## Abstract

**Introduction:** Clinical and population decision-making relies on the systematic evaluation of extensive regulatory evidence. The FDA drug reviews provide detailed information on clinical trial design, enrollment criteria, sample size, randomization, comparators, endpoints, and indications. However, extracting these data is resource-intensive and time-consuming. Generative Artificial Intelligence large language models (LLMs) may accelerate the extraction and synthesis of such information. This study compares the performance of three LLMs, ChatGPT-4o, Gemini 2.5 Pro, and DeepSeek R1, in extracting and synthesizing regulatory and clinical information using antibiotics approved for complicated urinary tract infections (cUTIs) between 2010 and 2025 as a case study.

**Methods:** LLM models were evaluated using general (short, direct) and detailed (structured, guidance-referencing) prompts across five domains including accuracy (precision and recall), explanation quality, error type (hallucination rate, misclassification, and omission), efficiency (response time, correct answers per second, and seconds per correct answer), and consistency with responses generated in duplicate runs. Two investigators independently reviewed outputs against FDA guidance, resolving discrepancies by consensus. Statistical analyses included χ², Wilcoxon, and Kruskal–Wallis tests with false discovery rate correction.

**Results:** Among 324 responses, accuracy differed significantly across models (χ², *p*<0.001) with Gemini 2.5 Pro achieving the highest accuracy (66.7%), followed by ChatGPT-4o (51.9%) and DeepSeek R1 (37.0%). General prompts outperformed detailed prompts (59.3% vs 44.4%; *p*=0.011). Gemini 2.5 Pro showed highest explanation quality and most consistent outputs, while ChatGPT-4o had shortest response times and highest efficiency. Hallucination was the most frequent error type across models.

**Conclusion:** LLMs showed variable capability in extracting regulatory and clinical trial information. Gemini 2.5 Pro showed the strongest overall performance, while ChatGPT-4o was faster but less accurate, and DeepSeek R1 underperformed across most domains. These findings highlight both the promise and limitations of LLMs in regulatory science and support complementary use alongside human review to streamline evidence synthesis.

**Author Summary:** Our research addresses a critical question in artificial intelligence for healthcare: how well do generative Generative Artificial Intelligence (GenAI) tools extract and synthesize regulatory and clinical information to inform decision-making? We assessed ChatGPT-4o, Gemini 2.5 Pro, and DeepSeek R1 performance in extracting and synthesizing information from regulatory documents and clinical trial data using all FDA approved antibiotics for the treatment of complicated urinary tract infections. We compared LLMs outputs directly with the original data sources. We assessed the models’ performance using both broad and detailed prompts across several areas, including accuracy of the information (precision and recall), quality of explanation, type of errors (hallucination, misclassification, and omission), efficiency and speed (response time, correct answers per second, and seconds per correct answer), and consistency of responses across repeated runs. The results suggest that while the models were generally fast and efficient extracting large volumes of information, they also produced errors and omissions that could limit their reliability. These findings highlight both the promise and the current limitations of GenAI, underscoring its potential value as a human supervised tool for safely supporting regulatory science and clinical decision-making.

## Introduction

The U.S. Food and Drug Administration (FDA) reviews preclinical and pivotal clinical trial information submitted in support of new drug approval applications to ensure compliance with regulatory standards and FDA guidance. Information generated through this review process forms the evidentiary basis for regulatory approval and is subsequently used by clinicians, health care organizations, payers, and patients to inform drug coverage, payment, and clinical practice decision-making. This information is distributed across multiple sources including FDA regulatory documents, medical, statistical, and clinical pharmacology reviews, and clinical trials.

Extracting, synthesizing, and interpreting information from FDA regulatory drug review documents is complex and typically requires interdisciplinary teams with expertise in clinical medicine, biostatistics, pharmacology, and regulatory science. As a result, the process is time-consuming, resource intensive, and difficult to scale, particularly as the volume and complexity of regulatory submissions continue to increase.

Recent advances in generative artificial intelligence (GenAI), particularly large language models (LLMs) such as ChatGPT, Gemini, and DeepSeek, have the potential to automate and augment the extraction, analysis, and summarization of complex regulatory drug information.(1–4) Recognizing this potential, the FDA is exploring LLM-based tools, including initiatives such as BERTox, to assist scientific reviewers and investigators in processing clinical trial information, with the goal of enhancing efficiency in the regulatory review process, reducing review time, and accelerating access to new therapies.(3,5,6)

Prior research has explored the use of LLMs to information extraction from structured regulatory documents, most notably FDA-approved drug labels.(7–9) However, FDA regulatory review documents differ substantially in structure, length, and complexity, often containing nuanced methodological details, interpretive commentary, and cross-referenced analyses. To date, no studies have systematically assessed the performance of LLMs in extracting, analyzing, and summarizing information from FDA regulatory review documents and clinical trials data, despite their central role in regulatory and downstream decision-making.

In this study, we address this knowledge gap by evaluating the accuracy, efficiency, and reliability of three state-of-the art LLMs, ChatGPT-4o, Gemini 2.5 Pro, and DeepSeek R1, in extracting, analyzing, and summarizing regulatory and clinical trial information. Using information from all FDA approved antibiotics for complicated urinary tract infections (cUTIs) since 2010, we provide a comparative assessment of LLM performance in a real-world regulatory context. Our findings inform the potential role of GenAI tools in supporting regulatory science and evidence synthesis in an era of increasing reliance on AI-assisted decision-making.

## Materials and Methods

We retrieved FDA drug reviews, guidance for the industry, and drug package insert data from publicly available sources including the Drugs@FDA databases(10–16) for the six antibiotics for cUTI (cefiderocol, imipenem/cilastatin/relebactam, plazomicin, meropenem/vaborbactam, ceftazidime/avibactam, and ceftolozane/tazobactam) approved by the FDA between 2010 and 2025. We purposefully selected cUTI due to the availability of FDA guidance for the industry to develop antibiotics for cUTI and the standardized clinical development and trial designs of antibiotics, which enabled a consistent comparison of clinical trial data and regulatory standards across antibiotics (16).

To assess the performance of ChatGPT-4o, Gemini 2.5 Pro, and DeepSeek R1, we used a combination of standardized information extraction metrics, including precision, recall, F1 score, and contextual attribute accuracy, explanation quality, error taxonomy, efficiency, and consistency across multiple runs (Table 1). These evaluation methods assessed language understanding, reasoning, and factual accuracy, as well as robustness under both expert review and stress-testing conditions designed to test model limitations.

**Table 1.** Summary of Evaluation Metrics, Definitions, Statistical Analyses, and Results across Large Language Models.

| **Metric** | **Definition** | **Statistical Test** | **Results** |
| --- | --- | --- | --- |
| Accuracy | Accuracy in identifying qualifiers and modifiers | Pearson's chi-square test: | $\chi^2(2) = 18.99, p < 0.001$ |
| Precision | % of correct extracted items | Pearson's chi-square test;<br>Two-sample proportion test (Pairwise) | Gemini 70%<br>ChatGPT 52%<br>DeepSeek 39% |
| Recall | % of correct items extracted | Pearson's chi-square test;<br>Two-sample proportion test (Pairwise) | Gemini 93.5%<br>ChatGPT 100%<br>DeepSeek 88% |
| F1 Score | Harmonic mean of precision and recall | - | Gemini 80%<br>ChatGPT 68%<br>DeepSeek 54% |
| Accuracy by prompt-type | Difference in accuracy between General vs Detailed prompts (and by LLM) | Pearson's chi-square test, pairwise risk differences with BH correction | Overall General prompts > Detailed prompts: 59.3% vs 44.4%, $\chi^2(1)=6.54, p=0.011$<br>**General prompts:** Gemini > DeepSeek ($\Delta=24.1$ pp; 95% CI 6.2–42.0; $p=0.011$)<br>**Detailed prompts:** Gemini > DeepSeek ($\Delta=35.2$ pp; 95% CI 17.7–52.7; BH-$p=0.0014$)<br>After FDR: Gemini > DeepSeek remained significant for both prompt types. Gemini vs ChatGPT was not significant. |
| Precision | % of correct extracted items | Pearson's chi-square test, Two-sample proportion tests (pairwise) | **General prompts:**<br>Gemini 75%<br>ChatGPT 61%<br>DeepSeek 54%<br>**Detailed prompts:**<br>Gemini 67%<br>ChatGPT 53%<br>DeepSeek 30% |

| Metric | Definition | Statistical Test | Results |
| --- | --- | --- | --- |
| Recall | % of relevant items successfully extracted | Pearson’s chi-square test,<br>Two-sample proportion tests<br>(pairwise) | **General prompts:**<br>Gemini 95%<br>ChatGPT 100%<br>DeepSeek 96%<br>**Detailed prompts:**<br>Gemini 100%<br>ChatGPT 100%<br>DeepSeek 88% |
| F1 Score | Harmonic mean of precision and recall (%) | — | **General prompts:**<br>Gemini 84%<br>ChatGPT 76%<br>DeepSeek 69%<br>**Detailed prompts:**<br>Gemini 81%<br>ChatGPT 69%<br>DeepSeek 45% |
| Explanation Quality | Manual scoring of factual accuracy, compliance, and formatting | Kruskal–Wallis test;<br>Wilcoxon rank-sum tests (Pairwise) | Kruskal–Wallis test; $\chi^2(2) = 23.48, p < 0.001$.<br>Wilcoxon rank-sum tests (pairwise)<br>Gemini > ChatGPT ($p = 0.014$)<br>Gemini > DeepSeek ($p < 0.001$)<br>ChatGPT > DeepSeek ($p = 0.047$) |
| Error Taxonomy |  |  |  |
| Hallucination Rate | % of outputs containing unsupported or fabricated content | Pearson's chi-square test | Pearson's chi-square test; ($\chi^2(2) = 10.38, p = 0.0056$).<br>ChatGPT > Gemini ($p = 0.0083$). |
| Misclassification | % of outputs with relevant content incorrectly labeled or categorized | Pearson's chi-square test | Pearson's chi-square test; LLMs ($\chi^2(2) = 2.10, p = 0.349$). |
| Omission | % of outputs missing required information | Fisher's exact test with Monte Carlo simulation (100,000 reps); BH-corrected pairwise Fisher tests; risk differences and 95% CIs | Fisher's exact test with Monte Carlo simulation (100,000 reps); ($p = 0.013$)<br>BH-corrected pairwise Fisher tests; risk differences and 95% CIs<br>ChatGPT < Gemini and DeepSeek ($p = 0.028$).<br>ChatGPT < Gemini ($\Delta = -14.3$ percentage points, 95% CI: $-25.9$ to $-2.7$)<br>ChatGPT < DeepSeek ($\Delta = -10.4$ percentage points, 95% CI: $-17.8$ to $-3.1$). |
| Efficiency |  |  |  |
| Response Time | Time (in seconds) from prompt submission to final model output display | Kruskal–Wallis test; Wilcoxon rank-sum tests (pairwise, BH-adjusted)<br>Median and IQRs | Kruskal–Wallis test; $\chi^2(2) = 205.8, p < 0.001$<br>Wilcoxon rank-sum tests (pairwise, BH-adjusted);<br>Median and IQRs:<br>ChatGPT: 20.7s (IQR: 18.3–24.1)<br>Gemini: 42.6s (IQR: 37.2–48.9)<br>DeepSeek: 45.6s (IQR: 37.2–58.9)<br>All pairwise differences were significant<br>ChatGPT > Gemini and DeepSeek<br>$p < 0.001$ |
|  | Correct answers per second (CPS) | Nonparametric bootstrap resampling (10,000 iterations) for means, CIs, and pairwise differences | Mean CPS (with bootstrap 95% CIs); |
|  |  |  | ChatGPT 0.0242 s<sup>-1</sup> (95% CI: 0.0196–0.0290)<br>Gemini 0.0154 s<sup>-1</sup> (95% CI: 0.0132–0.0175)<br>DeepSeek 0.0077 s<sup>-1</sup> (95% CI: 0.0053–0.0090) |
|  | Seconds per correct answer (SPC) | Nonparametric bootstrap resampling (10,000 iterations) for means, CIs, and pairwise differences | ChatGPT 41.7 s (95% CI: 34.5–51.1 s)<br>Gemini 65.4 s (95% CI: 57.1–75.5 s)<br>DeepSeek 144 s (95% CI = 111–190 s) |
| Consistency |  |  |  |
| Exact Match (EM) | Binary score whether a model output exactly matches reference (1 for exact match, 0 otherwise) | Pearson's chi-square test;<br>Two-sample proportion test (Pairwise, BH-corrected) | Pearson's chi-square test;<br>($\chi^2(2) = 18.99$, $p < 0.001$)<br>Pairwise two-sample proportion test<br>Gemini vs DeepSeek ($p < 0.001$)<br>Gemini vs ChatGPT ($p = 0.028$)<br>ChatGPT vs DeepSeek ($p = 0.028$) |

### Prompting Strategy

Two study investigators developed a predefined set of questions based on FDA guidance for the industry, including key drug information domains such as population characteristics, safety, and clinical trial design in pivotal Phase 2 and Phase 3 studies, and extracted data including trial design, randomization, blinding, active comparator, and trial type (noninferiority, superiority), inclusion and exclusion enrollment criteria, sample size, randomization procedures, comparators, primary and secondary endpoints, and statistical analysis considerations (S1 Appendix).

We manually extracted this information (reference answers) from the FDA review documents and the FDA-approved package inserts. We conducted a digital review using keyword searches (e.g., “population,” “safety,” “primary endpoint”), followed by manual review of hard copy documents. We compared reference answers to the FDA guidance to determine accuracy.

For LLM evaluation, we uploaded the corresponding PDF files, FDA drug reviews, industry guidance, and drug package inserts into each model. LLMs were prompted with two formats: general and detailed prompts. General prompts were brief, open-ended questions without specifying output format. Detailed prompts were structured, multi-part instructions referencing FDA guidance and requesting responses in a standardized checklist-type format. Examples of general and detailed prompts are included in (S2 Appendix). Each output was timestamped (in seconds), and metadata (model, prompt type, drug, and task) was recorded.

### Evaluation Metrics

Two study investigators independently assessed each response, using a predefined scoring protocol, and resolved discrepancies by consensus. Inter-rater reliability was assessed using percent agreement.

We assessed models (1) *accuracy*, defined as whether the LLMs identified qualifiers (e.g., drug name, clinical trial endpoint) and modifiers (e.g., dosage, frequency, duration, population subgroup specifications), (2) *precision* (i.e., percentage of extracted information that was correct), and (3) *recall* or the percentage of all correct items that the model successfully extracted (Table 1). We computed the *F1* score as the mean of precision and recall.

Additionally, we evaluated the *explanation quality* through human review, using a 5-point Likert scale (1 = minimal justification, 5= comprehensive and well-sourced) to rate the factual accuracy, clarity, and completeness of the model’s response.

We classified *incorrect responses* into one of three error types: (1) h*allucinations*, fabricated or unsupported claims, (2) m*isclassifications*, incorrect labeling of otherwise relevant content, and (3) o*missions or* failure to include required or expected information.

To assess each model efficiency, we recorded the time (in seconds) it took each model to generate a complete response using a stopwatch. We measure *response time* as the time interval from prompt submission to the model’s final output display. Then, we assessed two performance metrics: (1) Correct answers per second (CPS) defined as the number of correct responses divided by total time and (2) Seconds per correct answer (SPC) defined as the average time required to produce each correct answer. These metrics provide a composite measure of both speed and quality of responses.

To assess the reliability (consistency of the model’s responses), we repeated the entire evaluation four weeks after the initial run. We performed the first round in June 2025 and the second in July 2025. We measured reliability as a binary outcome: The binary score *exact match* captured whether the second output conveyed the same substantive content regardless of phrasing, formatting, or order and did not contradict the original response and *unmatched* captured whether the content differed in substance, omitted critical information, or had contradictions.

### Statistical Analysis

First, we compared the three LLMs using an omnibus test for each outcome, followed by pairwise comparisons adjusted for multiple testing using the Benjamini–Hochberg false discovery rate (FDR) method. We assessed prompt-type comparisons, general vs. detailed prompts, using Pearson’s chi-square tests on 2×2 tables, with risk differences and 95% confidence intervals (CI) estimated using prop.test() in R.

We assessed differences in *precision* and *accuracy* (binary outcomes) using Pearson’s chi-square test and pairwise comparisons using two-sample tests for proportions. Additionally, we assessed differences in *explanation quality* (ordinal) and *response time* (treated as skewed continuous data) using nonparametric methods. We used the Kruskal–Wallis test for overall comparisons across LLMs, followed by Wilcoxon rank-sum tests for pairwise contrasts. Skewness in response times was expected, as most responses were completed quickly with a few significant outliers.

For the *error taxonomy* among incorrect responses, we compared hallucination and misclassification rates using Chi-square tests. We used Monte Carlo simulation (100,000 replicates) for Fisher’s exact test to account for low expected cell counts in the omission error category. We applied the Benjamini–Hochberg correction for multiple comparisons and reported risk differences and corresponding 95% CIs. We also reported odds ratios with exact CIs for omission comparisons. Explanation quality scores had minimal missingness and no imputation was performed.

To assess differences in *efficiency* across LLMs, we used nonparametric bootstrap resampling (10,000 iterations) to estimate means, CIs, and pairwise statistically significant differences. We calculated both metrics, CPS and its inverse SPC for each LLM and estimated 95% CIs and pairwise contrasts using nonparametric bootstrap resampling (10,000 iterations).

To assess statistically significant differences across LLMs in *exact match*, we used Pearson’s chi-square test and pairwise comparison using **t**wo-sample proportion tests, with Benjamini–Hochberg (BH) correction. All statistical tests were two-sided with a significance threshold of α = 0.05. All analyses were conducted in R version 4.5.1 using RStudio.

## Results

In the period 2010-2025, the FDA approved six antibiotics for complicated urinary tract infections (cUTI), all of which remained on the market as of December 1, 2025. All three LLMs and study investigators successfully retrieved the requested information from FDA regulatory sources, drug reviews, industry guidance, and package inserts (S1 Appendix). The dataset comprised 324 model–prompt responses (6 drugs × 9 questions × 2 prompt types × 3 LLMs), balanced across models (n = 108 per LLM) and prompt types (162 general, 162 detailed).

### Accuracy

Accuracy varied significantly across LLMs (χ²(2) = 18.99, *p* < 0.001) (Table 2). Gemini 2.5 Pro achieved the highest accuracy at 66.7% (72/108), followed by ChatGPT-4o at 51.9% (56/108) and DeepSeek R1 at 37.0% (40/108).

**Table 2.** ChatGPT-4o, Gemini 2.5 Pro, and DeepSeek R1 Accuracy.

| Prompt type | Tool | Correct/Total | Accuracy %<br>[95% CI] | Omnibus<br>$\chi^2$ (df) | p-value | Pairwise contrasts (BH-adjusted) |
| --- | --- | --- | --- | --- | --- | --- |
| **Overall** | Gemini | 72 / 108 | 66.7 [57.3–74.8] | $\chi^2(2)=18.99$ | <0.001 | Gemini > ChatGPT ($p=0.040$);<br>ChatGPT > DeepSeek ($p=0.040$);<br>Gemini > DeepSeek ($p=7.3\times 10^{-5}$)— |
|  | ChatGPT-4o | 56 / 108 | 51.9 [42.5–61.0] |  |  |  |
|  | DeepSeek | 40 / 108 | 37.0 [28.5–46.4] |  |  |  |
| **General** | Gemini | 39 / 54 | 72.2 [59.1–82.4] | $\chi^2(2)=6.60$ | 0.037 | Gemini > DeepSeek ($\Delta=24.1$ pp;<br>95% CI 6.2–42.0; $p=0.011$) |
|  | ChatGPT-4o | 31 / 54 | 57.4 [44.2–69.7] |  |  |  |
|  | DeepSeek | 26 / 54 | 48.1 [35.4–61.1] |  |  |  |
| **Detailed** | Gemini | 33 / 54 | 61.1 [47.8–73.0] | $\chi^2(2)=13.65$ | 0.001 | Gemini > DeepSeek ($\Delta=35.2$ pp;<br>95% CI 17.7–52.7; $p=0.0014$); other contrasts ns |
|  | ChatGPT-4o | 25 / 54 | 46.3 [33.7–59.4] |  |  |  |
|  | DeepSeek | 14 / 54 | 25.9 [16.1–38.9] |  |  |  |

BH: Benjamini–Hochberg false discovery rate correction; ns: Not significant
Note: across LLMs, accuracy was higher for General vs Detailed prompts (59.3% vs 44.4%; $\chi^2(1) = 6.54$ , $p = 0.011$ ).

LLM accuracy varied by prompt type. General prompts consistently outperformed detailed prompts (59.3% vs 44.4%; χ²(1) = 6.54, p = 0.011). For general prompts, Gemini 2.5 Pro accuracy was higher than DeepSeek R1 (72.2% vs 48.1%; risk difference [Δ] = 24.1 percentage points, 95% CI: 6.2–42.0; *p* = 0.011). Accuracy also differed across LLMs for detailed prompts (χ²(2)=13.65, *p* = 0.001). Gemini 2.5 Pro also outperformed DeepSeek R1 (61.1% vs 25.9%; Δ = 35.2 percentage points, 95% CI: 17.7–52.7; Benjamini–Hochberg correction *p* = 0.0014) for detailed prompts. After FDR correction, only Gemini’s accuracy remained significantly greater than DeepSeek’s for both prompt types general and detailed. The difference between Gemini 2.5 Pro and ChatGPT-4o was not statistically significant (Fig 1).

**Fig 1.**
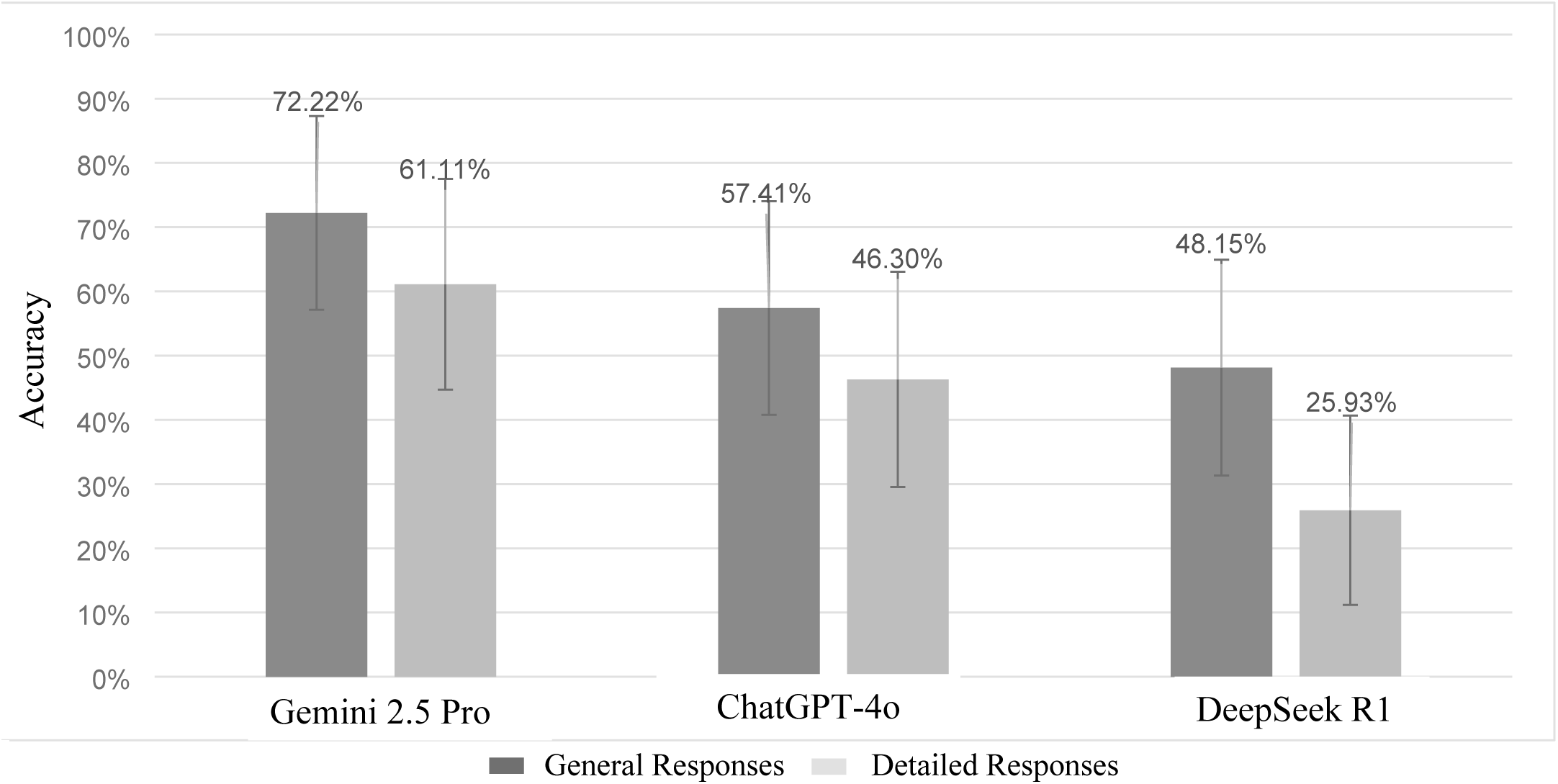
LLMs Accuracy by Prompt Type: General and Detail Prompts.

Accuracy: Proportion of correct responses. Bars show point estimates. Error bars represent Wilson 95% Confidence Intervals.

Among the three LLMs, Gemini achieved the highest F1 score (0.80) with high precision (70%) and high recall (93.5%) (Table 1). ChatGPT consistently achieved perfect recall (100%), but lower precision (52%), resulting in a lower F1 score (0.68). DeepSeek showed the lowest precision (39%) and recall (88%) and thus, the lowest F1 score (0.54). General prompts consistently outperformed detailed prompts across models. Gemini 2.5 Pro achieved high precision and recall followed by ChatGPT-4o. For detailed prompts, DeepSeek R1 achieved the lowest precision and recall, which dropped the F1 score to 0.45.

### Explanation Quality

Explanation quality scores also varied significantly by LLM (Kruskal–Wallis χ²(2) = 23.48, *p* < 0.001). Mean scores (1–5 scale) were highest for Gemini 2.5 Pro (mean = 3.72), followed by ChatGPT-4o (3.22), and DeepSeek R1 (2.89). Pairwise Wilcoxon tests with Benjamini–Hochberg correction showed that Gemini 2.5 Pro outperformed both ChatGPT-4o (*p* = 0.014) and DeepSeek R1 (*p* < 0.001) and that ChatGPT-4o outperformed DeepSeek R1 (*p* = 0.047) (Fig 2).

**Fig 2.**
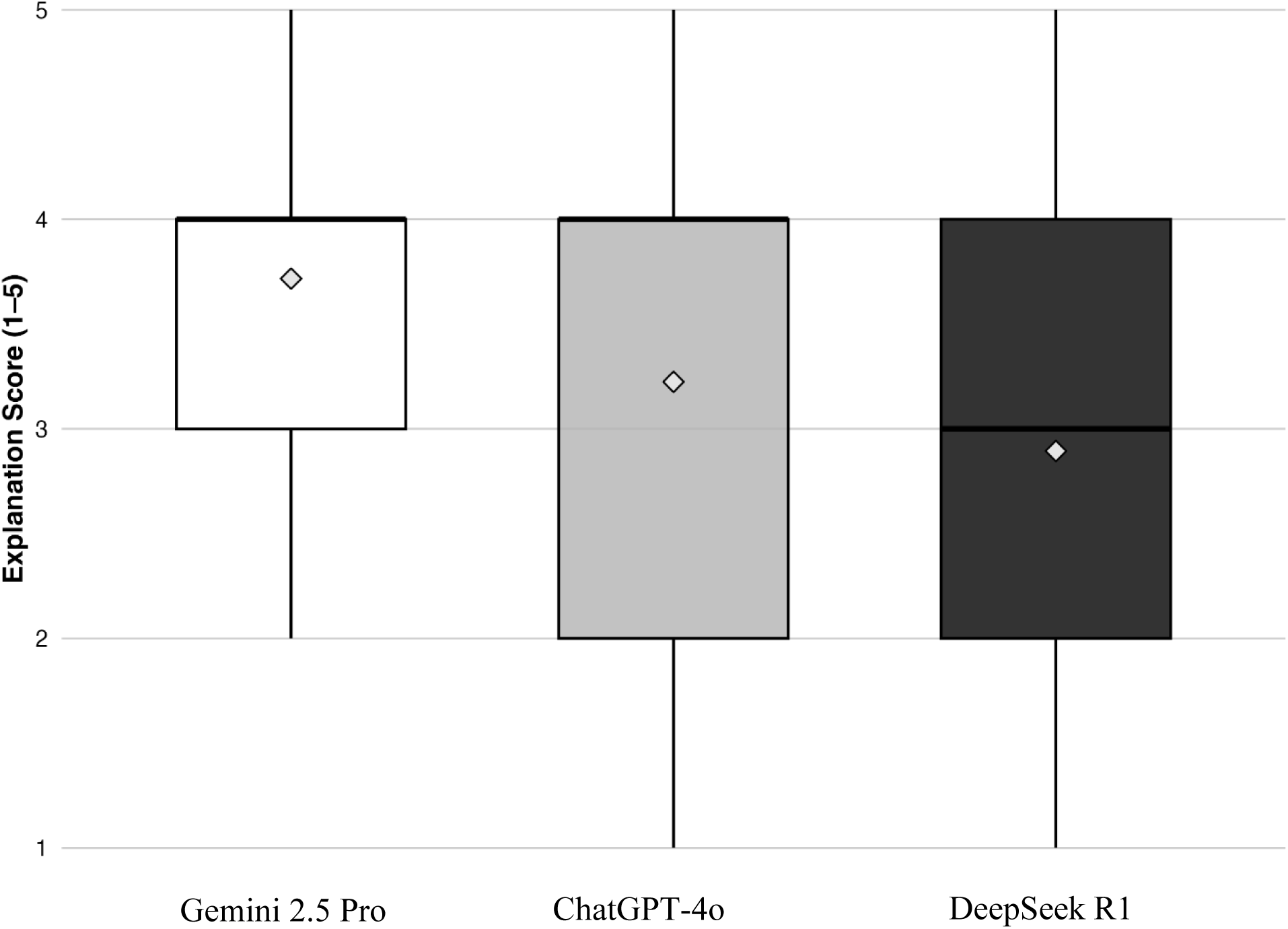
Quality Scores by Generative Artificial Intelligence Large Language Model.

Boxes show the interquartile range (IQR) of explanation scores. The horizontal line within the box indicates the median and the diamond symbol indicates the mean.

### Error Taxonomy

Among incorrect responses, the hallucination rate differed significantly by model (χ²(2) = 10.38, *p* = 0.0056) (Table 1). ChatGPT-4o had the highest hallucination rate at 74.5% (38/51), followed by DeepSeek R1 at 61.2% (41/67), and Gemini 2.5 Pro at 40.0% (14/35). Pairwise comparisons showed that ChatGPT-4o had significantly more hallucinations than Gemini 2.5 Pro (Benjamini–Hochberg adjusted *p* = 0.0083). Pairwise analysis was not statistically significant after Benjamini–Hochberg correction for multiple testing (ChatGPT-4o vs DeepSeek R1, *p* = 0.128; DeepSeek R1 vs Gemini 2.5 Pro, *p* = 0.062).

Conversely, *misclassification* rates did not differ significantly across LLMs (χ²(2) = 2.10, *p* = 0.349). The proportion of misclassified responses was 57.1% for Gemini 2.5 Pro (20 of 35), 43.1% for ChatGPT-4o (22 of 51), and 43.3% for DeepSeek R1 (29 of 67).

*Omission* errors, while relatively infrequent, did vary by model (Fisher’s exact test with Monte Carlo simulation, *p* = 0.013). ChatGPT-4o output had no omissions (0/51) compared to omission rates of 14.3% (5/35) for Gemini 2.5 Pro and 10.4% (7/67) for DeepSeek. Pairwise Fisher tests with Benjamini–Hochberg correction showed omission rates were significantly lower for ChatGPT-4o compared to both Gemini 2.5 Pro (*p* = 0.028) and DeepSeek (*p* = 0.028). The difference between Gemini and DeepSeek was not statistically significant (*p* = 0.747).

Estimated risk differences confirmed these findings. ChatGPT-4o had substantially fewer omissions than Gemini (Δ = −14.3 percentage points, 95% CI: −25.9 to −2.7) and DeepSeek (Δ = −10.4 percentage points, 95% CI: −17.8 to −3.1).

### Efficiency

Response time differed significantly across LLMs (Kruskal–Wallis χ²(2) = 205.8, *p* < 0.001). ChatGPT was the fastest model (median 20.7s; IQR: 18.3–24.1) compared to Gemini (median 42.6s; IQR: 37.2–48.9) and DeepSeek (median 45.6s; IQR: 37.2–58.9) (Table 3). All pairwise differences were significant (ChatGPT-4o vs Gemini 2.5 Pro and DeepSeek R1, both *p* < 0.001; Gemini 2.5 Pro vs DeepSeek R1, *p* = 0.019). The difference in response time was not statistically significant between general and detailed prompts overall (*p* = 0.999) or across models (e.g., DeepSeek, *p* = 0.140) (Fig 3).

**Table 3.**
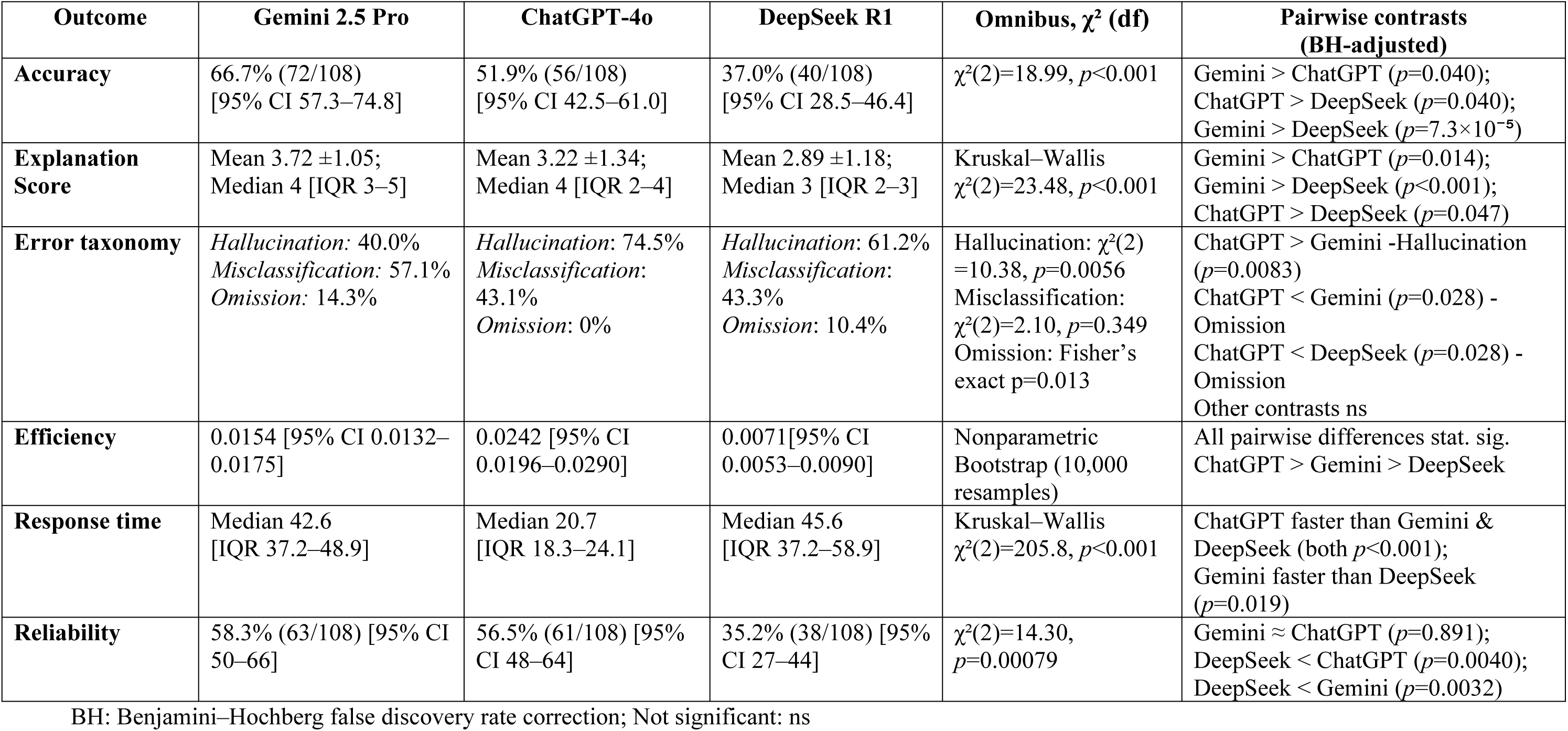
ChatGPT-4o, Gemini 2.5 Pro, and DeepSeek R1 Performance.

**Fig 3.**
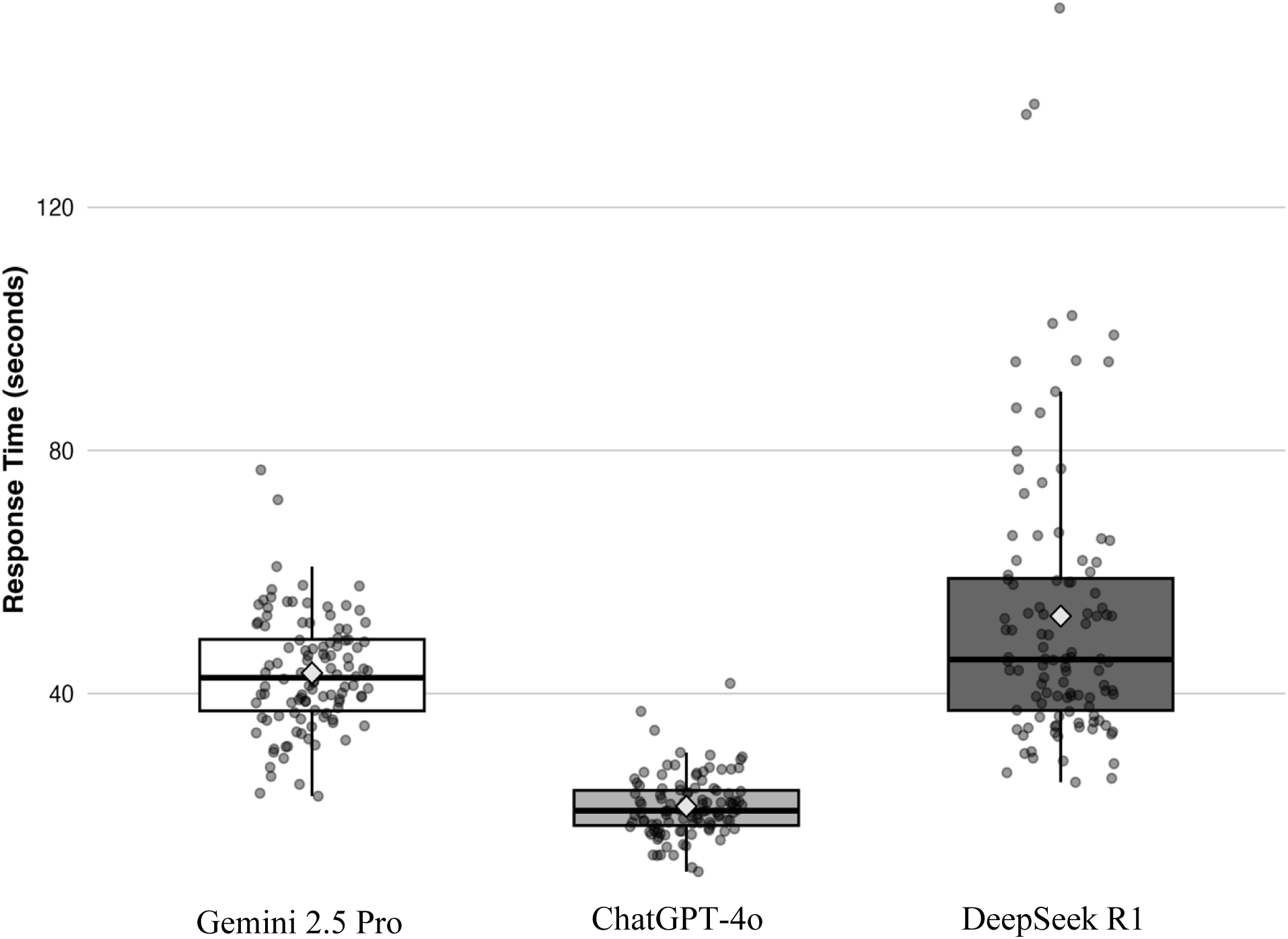
Response Time (in seconds) by Generative Artificial Intelligence Large Language Model.

Response time distributions (in seconds) by LLM. Boxes show medians and IQRs; whiskers extend to 1.5×IQR; diamonds mark means; points represent individual responses.

The number of correct answers per second (CPS) and seconds per correct answer (SPC) varied by model. The mean CPS (with bootstrap 95% CIs) was 0.0242 s⁻¹ (95% CI: 0.0196–0.0290) for ChatGPT-4o, 0.0154 s⁻¹ (95% CI: 0.0132–0.0175) for Gemini 2.5 Pro, and 0.0077 s⁻¹ (95% CI: 0.0053–0.0090) for DeepSeek R1 (Table 1). All pairwise CPS differences were statistically significant and large enough to affect real-world model’s performance. The difference in CPS was statistically significant among models including ChatGPT-4o vs. Gemini (0.00883s; 95% CI: 0.00386–0.0141), ChatGPT-4o vs. DeepSeek (0.0171s; 95% CI: 0.0121–0.0223), and Gemini vs. DeepSeek (0.00831; 95% CI: 0.00544–0.0111).

CPS ratios showed a similar pattern. ChatGPT-4o was 1.58 times more efficient than Gemini 2.5 Pro (95% CI: 1.24–1.99) and 3.49 times more efficient than DeepSeek R1 (95% CI: 2.48–4.87). Gemini was also more efficient than DeepSeek, with a CPS ratio of 2.22 (95% CI: 1.64–3.01).

Findings for SPC are consistent with CPS results. On average, ChatGPT-4o required approximately 41 seconds per correct answer, compared to 65 seconds for Gemini 2.5 Pro and 144 seconds for DeepSeek R1 (Fig 4).

**Fig 4.**
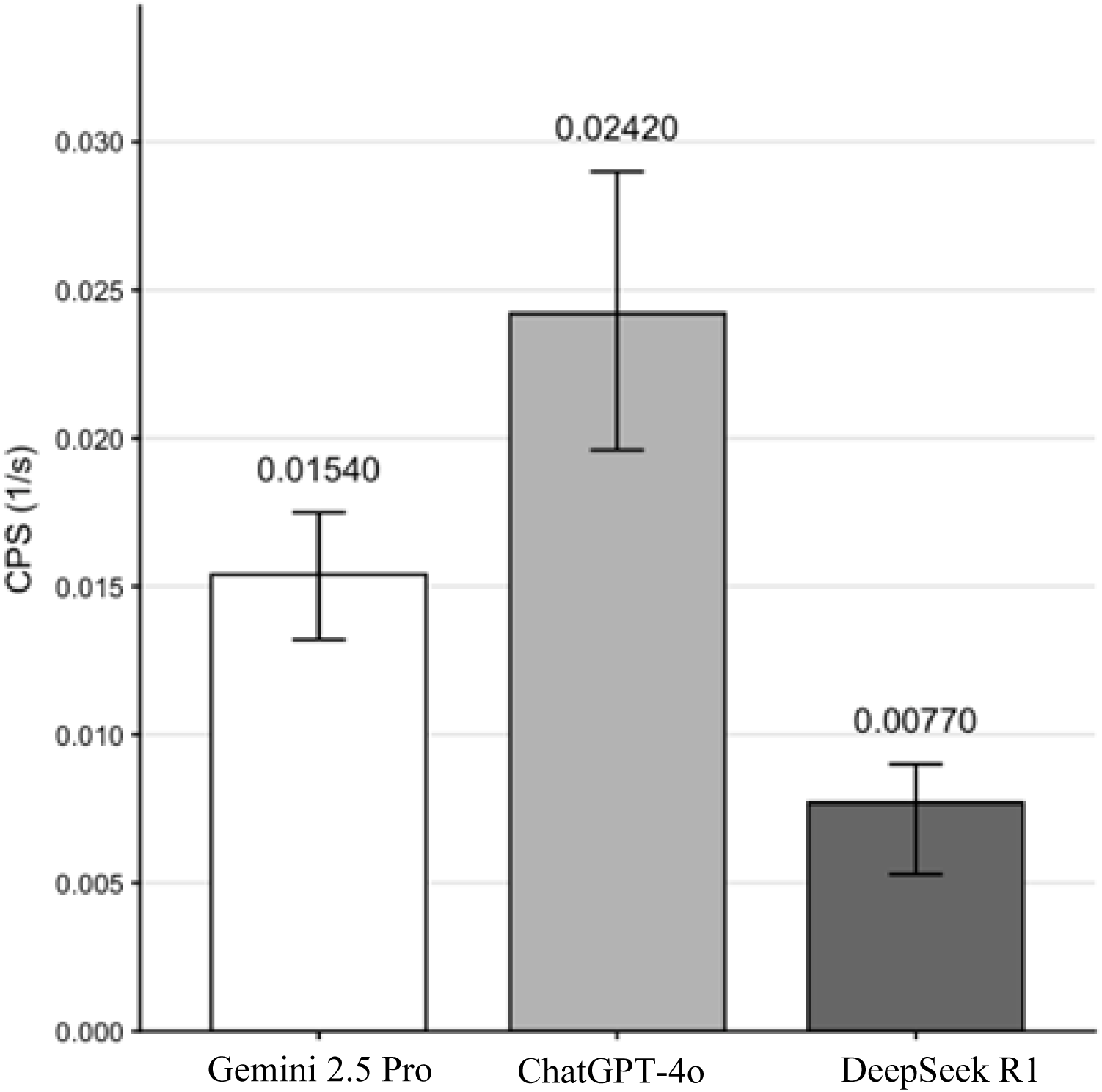
Generative Artificial Intelligence Large Language Model Efficiency.

Bars represent mean CPS (1/s) with error bars indicating the standard error of the mean (SEM).

### Consistency

The mean *exact match* rate across all models (N = 324) was 0.519 with significant differences in the *exact match* by model (χ²(2) = 18.99, *p* < 0.001) (Table 2). Gemini achieved the highest *exact match* rate (0.667, 95% CI 0.573–0.748), followed by ChatGPT (0.519, 95% CI 0.425–0.610) and DeepSeek (0.370, 95% CI 0.285–0.464). Each model was evaluated on 108 outputs, yielding 72 exact matches for Gemini, 56 for ChatGPT, and 40 for DeepSeek. Pairwise two-sample proportion tests with Benjamini–Hochberg correction confirmed that all pairwise differences were statistically significant (Table 1), suggesting significant performance differences across models.

### Reliability

Consistency across repeated runs also differed significantly by model (χ²(2) = 14.30, *p* = 0.00079). Gemini 2.5 Pro and ChatGPT-4o showed comparable consistency (58.3% [63/108] and 56.5% [61/108], respectively; p = 0.891); whereas, DeepSeek R1 was significantly less consistent at 35.2% (38/108) (Table 3). Pairwise comparisons with Benjamini–Hochberg correction also showed DeepSeek’s significantly lower match rate compared to both Gemini 2.5 Pro (*p* = 0.0032) and ChatGPT-4o (*p* = 0.0040) (Fig 5).

**Fig 5.**
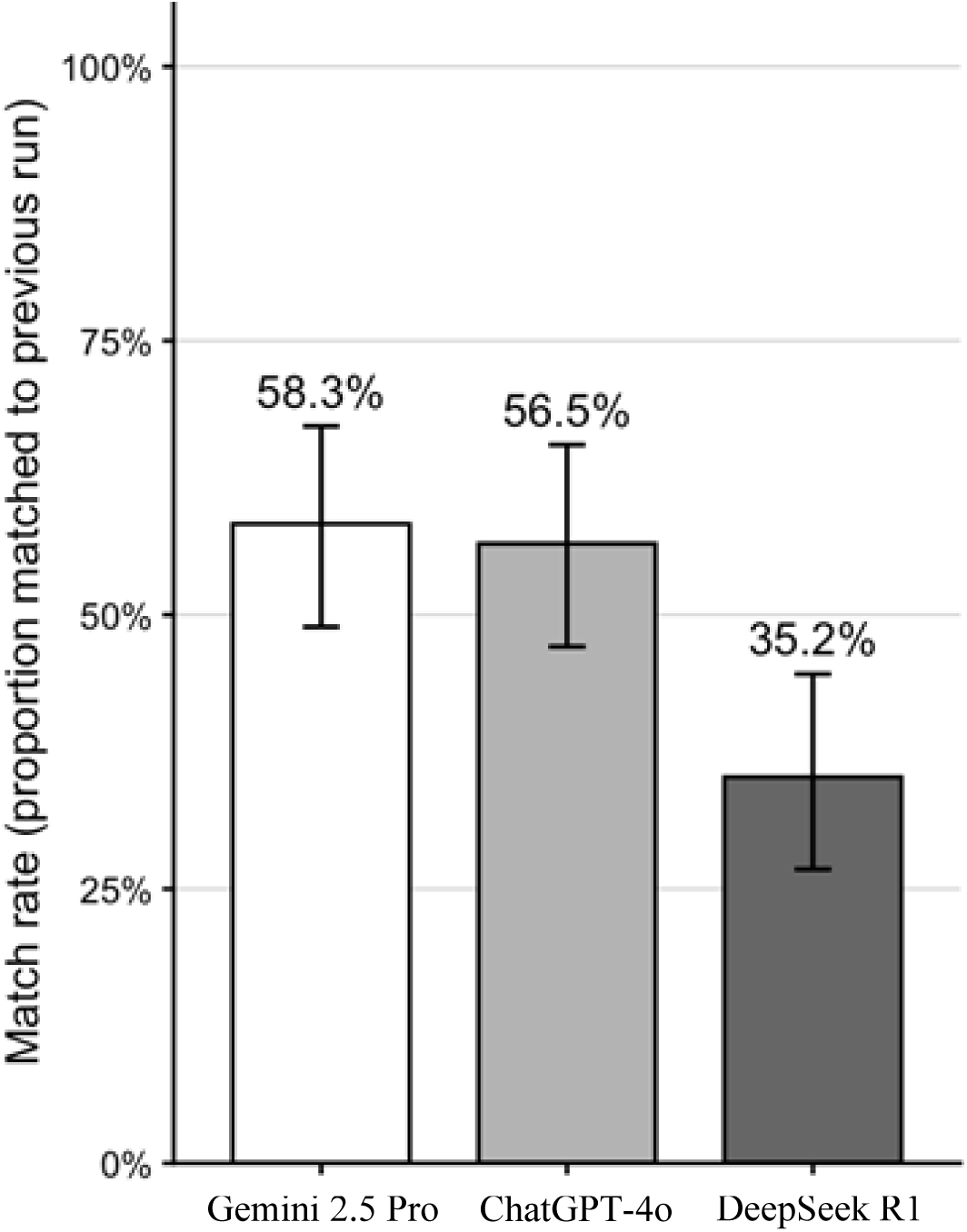
Consistency of Responses across Repeated Runs.

Bars represent the mean proportion of responses matching the previous run, with error bars indicating the standard error of the mean (SEM)

## Discussion

Generative Artificial Intelligence large language models have the potential to enhance retrieval and processing of regulatory and biomedical data, thereby informing decision-making by regulators, payers, healthcare providers, and patients.(4) In this comparative analysis assessing the accuracy, efficiency, and reliability of ChatGPT-4o, Gemini 2.5 Pro, and DeepSeek R1 in performing regulatory and clinical trial related tasks, we found that overall LLM performance varied significantly across models, with the highest performance observed for Gemini 2.5 Pro and the lowest for DeepSeek R1.

Accuracy lies at the core of LLMs reliability in regulatory and clinical applications. This study found that general prompts consistently outperformed detailed prompts across models, with no statistically significant differences between Gemini 2.5 Pro and ChatGPT-4o; both models achieved greater accuracy than DeepSeek R1 for both general and detailed prompt types. Gemini 2.5 Pro achieved the highest F1 score, with the greatest precision and recall. While ChatGPT-4o consistently achieved near perfect recall, it also had a significantly lower precision suggesting inclusion of excessive or inaccurate information. DeepSeek R1 performed the poorest, with the lowest scores in both precision and recall.

Explanation quality also varied by model, being highest for Gemini 2.5 Pro, followed by ChatGPT-4o, and lowest for DeepSeek R1. Importantly, explanation quality did not necessarily correlate with accuracy, but instead provided additional helpful elements such as citations, definitions, and clinical trial details which made manual verification of the responses easier.

LLMs are known to produce inaccurate responses or fabricated information (“hallucinations”). While prompt constraints and retrieval grounding may help prevent taxonomy-related errors, they do not fully eliminate hallucinations.(17) We found that ChatGPT-4o exhibited the highest hallucination rate, followed by DeepSeek R1 and Gemini 2.5 Pro. However, ChatGPT-4o rarely omitted required elements, consistently including all relevant information. Conversely, Gemini’s incorrect answers were less likely to contain hallucinations but more prone to omitting key details. We did not find any significant differences in the misclassification rates among the models. These findings suggest that Gemini 2.5 Pro can provide the most reliable balance of precision and completeness, whereas ChatGPT provides the greatest comprehensiveness, albeit at the expense of precision. Investigators should make sure Gemini 2.5 Pro output is appropriately sourced, and not to interpret GPT-4o thorough responses as accurate.

In terms of efficiency, ChatGPT-4o had the fastest response time and DeepSeek R1 the slowest. ChatGPT-4o also achieved the highest number of correct answers per unit time and the lowest time per correct answer, suggesting its utility in time-sensitive applications.

LLMs performance extracting information varies widely depending on the model, prompt design, and task complexity.(18,19) Prompt engineering is a critical determinant of model accuracy ineffective prompts can result in inaccurate outputs.(20) We found that short, general prompts generated more accurate responses than lengthy, rule-heavy detailed prompts. General prompts allowed the model to focus on a single task (e.g., “Did ≥30% have pyelonephritis?”) without needing to juggle excessive formatting constrains. These findings align with the *“*Lost in the Middle” phenomenon, where model performance declines when relevant information is embedded in long prompts or context blocks.(21) Similarly, in our study, detailed prompts broke down the task and required structured output with the models presenting the information in a specific template rather than searching and extracting the facts directly from the source documents. Our findings are consistent with recent work showing that LLMs with extended reasoning chains can increase hallucination, especially when uncertainty is not aligned with factual accuracy.(22)

Our findings regarding accuracy, recall, and precision over identical queries repeated over time are also consistent with prior work.(23) While LLMs may be highly consistent in tasks like binary classification and sentiment analysis(24), they can still produce incorrect responses and fabricated references(25,26) underscoring the need for human oversight particularly in the clinical and regulatory fields.

In healthcare, ChatGPT has been used for tasks such as extracting cancer treatment data and generating real-time responses for patients.(27,28) LLMs have also been applied to extract cognitive assessment scores, such as the Mini-Mental State Examination (MMSE) and Clinical Dementia Rating (CDR), from unstructured clinical notes for patients with Alzheimer’s disease and related dementias, achieving accurate results.(29) AI models have also been used to identify drug-disease associations and prevent medication errors, suggesting that LLMs may be a helpful tool in the search for drug-related information.(6,30) Specialized models, such as BioBERT and AskFDALabel, have outperformed conventional natural language processing approaches in extracting drug-drug interactions from unstructured biomedical texts.(27,31,32) The AskFDALabel tool, in particular, has shown effectiveness in extracting adverse events data from FDA drug labels, streamlining drug safety studies.(7) These applications have the potential to significantly transform how healthcare providers and patients access and interact with their biomedical data.

To our knowledge, this is the first study to systematically assess LLMs performance in extracting regulatory and clinical trial information, directly comparing prompt types and assessing multiple performance metrics. By measuring accuracy, explanation quality, efficiency, consistency, and error taxonomy, this study provides a comprehensive, multidimensional evaluation of LLMs performance – a critical step toward validating these tools for use in regulatory and clinical decision-making.

### Limitations

Our study used approved for cUTI, which may limit the generalizability of findings to other therapeutic classes or indications. Additionally, LLM performance may vary over time, as the same prompt can yield different outputs such as changes in accuracy, speed, or error types, before and after model updates.(33) We evaluated LLMs performance using prespecified prompt templates. Since LLMs are sensitive to prompt wording and instruction order, our findings should be interpreted as prompt-dependent and may vary under different prompt designs.(21, 34, 35) We defined response consistency at the content level, based on agreement on prespecified key clinical or regulatory elements, rather than requiring verbatim matches. Our approach reflects how regulatory reviewers assess consistency and alignment in practice. Despite the high accuracy observed for some models, human validation remains essential to identify and mitigate errors, hallucinations, and misinterpretations.(36)

## Conclusions

In this comparative evaluation of three state-of-the-art LLMs -ChatGPT-4o, Gemini 2.5 Pro, and DeepSeek R1- for extracting, analyzing and synthesizing regulatory and clinical trial data for all FDA approved antibiotics for cUTI performance varied significantly by model and prompt type. Gemini 2.5 Pro showed the highest overall accuracy, explanation quality, and F1 score, while ChatGPT-4o offered the most comprehensive outputs with superior recall, fastest response times, and highest efficiency per correct answer. DeepSeek R1 consistently underperformed across all metrics. General prompts outperformed detailed prompts, suggesting that task simplification improves LLM output reliability. ChatGPT-4o exhibited a higher hallucination rate, but it rarely omitted relevant content, whereas Gemini 2.5 Pro produced more concise and accurate answers but with occasional omissions. These findings underscore the evolving potential of LLMs to support regulatory and clinical data extraction and synthesis, while also highlighting the need for prompt optimization, model-specific validation, and continued human oversight.

## Funding/Support

The Laura and John Arnold Foundation provided grant support (ID # 25-14625). The authors thank the Deanship of Graduate Studies and Scientific Research at Taif University for covering the journal publication fee.

## Role of the Funder/Sponsor

The Laura and John Arnold Foundation had no role in the study design or conduct of the study; collection, management, analysis, and interpretation of the data; preparation, review, or approval of the manuscript; and decision to submit the manuscript for publication. Taif University had no role in the conduct of the research, manuscript preparation, or publication decision.

## Conflicts of Interest

None declared.

## Data Availability

The data used for the study are available at the FDA website Drugs@FDA. https://www.accessdata.fda.gov/scripts/cder/daf/index.cfm

## Notes

### Competing Interest Statement

The authors have declared no competing interest.

### Funding Statement

Yes

